# Acceptability of and confidence in novel tuberculosis vaccines among South African adults living with HIV

**DOI:** 10.64898/2026.07.10.26357768

**Authors:** Alexa Giovanatti, Nsika Sithole, Indira Govender, Heidi J. Larson, Limakatso Lebina, Paul K. Drain, Connie Celum, R. Scott McClelland, Jennifer M. Ross, Alison D. Grant, Adrienne E. Shapiro

## Abstract

**Objectives:** Several novel tuberculosis vaccines (NTVs) are being evaluated in clinical trials. Understanding the perception of acceptability and confidence in NTVs will inform implementation strategies.

**Methods:** We conducted a cross-sectional survey to assess acceptability in and confidence of NTVs among adult persons with HIV (PWH) at two clinics in KwaZulu-Natal province, South Africa. We evaluated associations between acceptability and sociodemographic factors.

**Results:** Among 225 PWH, 112 (50%) responded “Definitely yes, as soon as available”, 86 (38%) “Definitely yes, but wait ≥6 months to receive it” and 27 (12%) “Unsure, leaning yes, any timeline” to an NTV. Acceptability was associated with perception of tuberculosis’s (TB’s) importance in the community, perceived risk of contracting TB, personal history of TB, not currently living with someone with TB, incompatibility of vaccines with religion, and unemployment. Most participants were confident in the vaccine’s potential safety (110, 93%) and effectiveness (221, 98%). Participants preferred to receive information from government, community, and health entities, or internet, and vaccination at community settings over public health facilities.

**Conclusions:** NTV acceptability was high amongst these PWH, and they preferred community-based delivery models. Our findings may inform strategies to increase implementation of NTVs among PWH in South Africa.

## Introduction

Tuberculosis (TB) remains the leading infectious cause of death worldwide with 1.23 million deaths in 2024, disproportionately impacting people in low- and middle-income countries (LMIC) [1]. The only licensed TB vaccine is the Bacillus Calmette-Guérin (BCG), developed in 1921, which primarily serves to prevent severe forms of TB in children but does not prevent TB in adolescents or adults [2]. A NTV for adolescents and adults in 105 LMIC has been projected to prevent 4.6-8.5 million deaths by 2050 and increase gross domestic product by US$1.6 trillion by 2080, making this a priority investment for global stakeholders [3]. Galvanized by the World Health Organization’s (WHO) End TB Strategy launched in 2014, several promising vaccine candidates to prevent TB in adults and adolescents are now being evaluated in clinical trials [4].

Despite the demonstrated need for novel tuberculosis vaccines (NTVs), understanding of the attitudes of the intended recipients towards a NTV is extremely limited. This evidence gap is particularly important in high TB prevalence settings like South Africa and among people with HIV (PWH) who could benefit the most from a NTV.

Vaccine hesitancy is a critical public health concern with rising vaccine hesitancy following the COVID-19 pandemic and increased politicization of vaccines worldwide [5]. One study found that 25.6% of South Africans reported that experiences with the COVID-19 vaccine process made them less likely to get vaccinated in the future [6]. In a global survey of 23 countries during June-July 2022, South Africa reported the highest COVID vaccine hesitancy at 52.1% [7]. In preparing for introducing a NTV, vaccine acceptability studies are critical to identify drivers of uptake which can in turn inform demand creation strategies, motivate key stakeholders to execute vaccine programs, and support effective and equitable vaccine rollout [8,9]. Acceptability of a NTV in South Africa was hypothesized be similar to acceptance of overall vaccines which is about 75.5% in South Africa [10]. Acceptability studies of COVID-19, HPV, and childhood immunizations in South Africa found that vaccine acceptability was inversely associated with young age [11–14], higher perceived risk of disease [15], concerns about vaccine safety or efficacy [11,13–16], and social media use for health information [11,12]. The relationship of these demographic and behavioral factors to NTV acceptability in South Africa has not been assessed.

To address this gap, ambulatory adult PWH in South Africa were surveyed to assess their acceptance and confidence in NTVs. This sampled cohort represents a key recipient population for a NTV since the burden of TB, multidrug-resistant (MDR) TB, HIV-associated TB, and TB mortality rates among PWH in South Africa are among the highest worldwide [2,17].

## Methods

### Study design

The NTV acceptability survey (VAS) was a cross-sectional, observational study nested within DROP-TB; a prospective cohort study evaluating the use of tools including a novel urine LAM assay for TB diagnosis for PWH at the time of HIV care entry, with a 12-month follow-up period to assess TB preventive therapy uptake and incident TB [18]. Written consent was required for participation. At study enrollment, participants underwent a baseline clinical exam and provided samples for routine and investigative testing including HIV and TB labs. The baseline evaluation included standardized questions about demographic and clinical history including personal and family TB history and COVID-19 vaccination status. Thereafter, participants attended quarterly follow-up visits for one year during which the VAS was administered once per participant.

### Setting

DROP-TB and the VASs were conducted at two public Department of Health primary care clinics in the uMkhanyakude district in KwaZulu-Natal province, South Africa. One was a large, peri-urban clinic and the other small and rural. DROP-TB study enrollment took place between December 2021 and May 2024 and the VAS was conducted beginning June 2024 and ending October 2024.

### Participants

All DROP-TB participants remaining in follow-up in June 2024 were invited to participate in the VAS. DROP-TB eligibility criteria included: able and willing to provide written informed consent for study procedures; age >= 18 years; presenting to the study clinics for HIV care; a positive HIV test documented in the medical chart; initiating (naive) or reinitiating antiretroviral therapy (ART) after at least 90 days; had not received TB therapy in the past 90 days; and residing in the study area. In 2022, DROP-TB enrollment was augmented by co-enrolling participants in the Africa Health Research Institute Household Contact Study (AHCOS) [19]. Co-enrolled AHCOS participants included adult PWH already on ART, with a new positive test result for TB but not yet on TB treatment.

### TB Acceptability Survey Instrument

The VAS was adapted from the Vaccine Confidence Project’s (VCP) Vaccine Confidence Index (VCI), which explores individual perception of the importance, safety, effectiveness, and compatibility of vaccines with their beliefs. The VCI questionnaire has been used in large-scale, global monitoring of vaccine confidence since 2015 [20]. The VAS retained the original VCI domains and included additional items assessing NTV confidence and acceptability, perception of TB disease compared to other endemic infectious diseases in South Africa, and vaccine platform preferences (Supplement).

Vaccine confidence and acceptability were defined as follows:

**Vaccine confidence (WHO definition)**: the belief that vaccines are effective, safe, and part of a trustworthy medical system. Low vaccine confidence is distinct from, but may contribute to, vaccine hesitancy [21].

**Vaccine acceptability:** the opposite of vaccine hesitancy, which the WHO defines as the delay in acceptance or refusal of vaccination despite availability of vaccination services [21].

Accordingly, vaccine acceptability was represented by the composite of two questions addressing vaccine willingness and preferred timing for take a vaccine. (Willingness: *Scientists are developing new vaccines to prevent tuberculosis (TB). Several TB vaccines are being tested in clinical trials right now. If a vaccine is shown to be successful in a clinical trial…how willing would you be to accept the vaccine;* Time of receipt: *How soon would you receive a tuberculosis (TB) vaccine that is approved for use in South Africa and available to you personally).* Vaccine confidence was derived from the questions about NTV safety and effectiveness. Participants’ trust in the medical system was not directly assessed, but participants were asked about their preferred source of health information and location to receive a vaccine.

### Data collection

At the time the VAS was introduced, participants were reminded of the voluntary nature and confidentiality of <u>the</u> survey <u>and</u> their right to stop answering or decline to answer at any point. A study nurse administered the survey in the participant’s preferred language (English or isiZulu) in person or by telephone. Responses were recorded electronically on a tablet in a REDCap database by the study nurse.

### Data analysis

R version 2024.09.1+394 software was used to perform statistical analysis (Foundation for Statistical Computing, Vienna, Austria). Descriptive statistics were used to summarize all variables. Kruskal-Wallis analyses were used due to the ordinal nature of the outcome, non-normal distribution of data, and small sample size. In this exploratory analysis, to avoid multiple comparisons, a limited set of variables were selected based on existing literature on the acceptability of COVID, HPV, and childhood immunizations. For several variables, response categories with small counts were combined where appropriate to ensure sufficient sample size for statistical testing. For variables with more than two categories that were significant, post-hoc Dunn’s testing was performed to examine pairwise comparisons within that variable. An alpha-value of 0.05 was considered significant.

## Results

A total of 225 participants were surveyed, 191 from DROP-TB and 34 from AHCOS. Of these, 129 (57%) were female and 225 (100%) were Black African. Median age was 34 years (IQR 27-42) and median baseline CD4 count was 338 cells/ul (IQR: 179-514). Full demographic characteristics are described in Table 1. In the study cohort, 64 (28%) reported a personal history of past or current TB, 92 (41%) were living with someone with TB at the time of survey, 216 (96%) had ever lived someone with TB, and 173 (77%) had never received a COVID-19 vaccine.

**Table 1:**
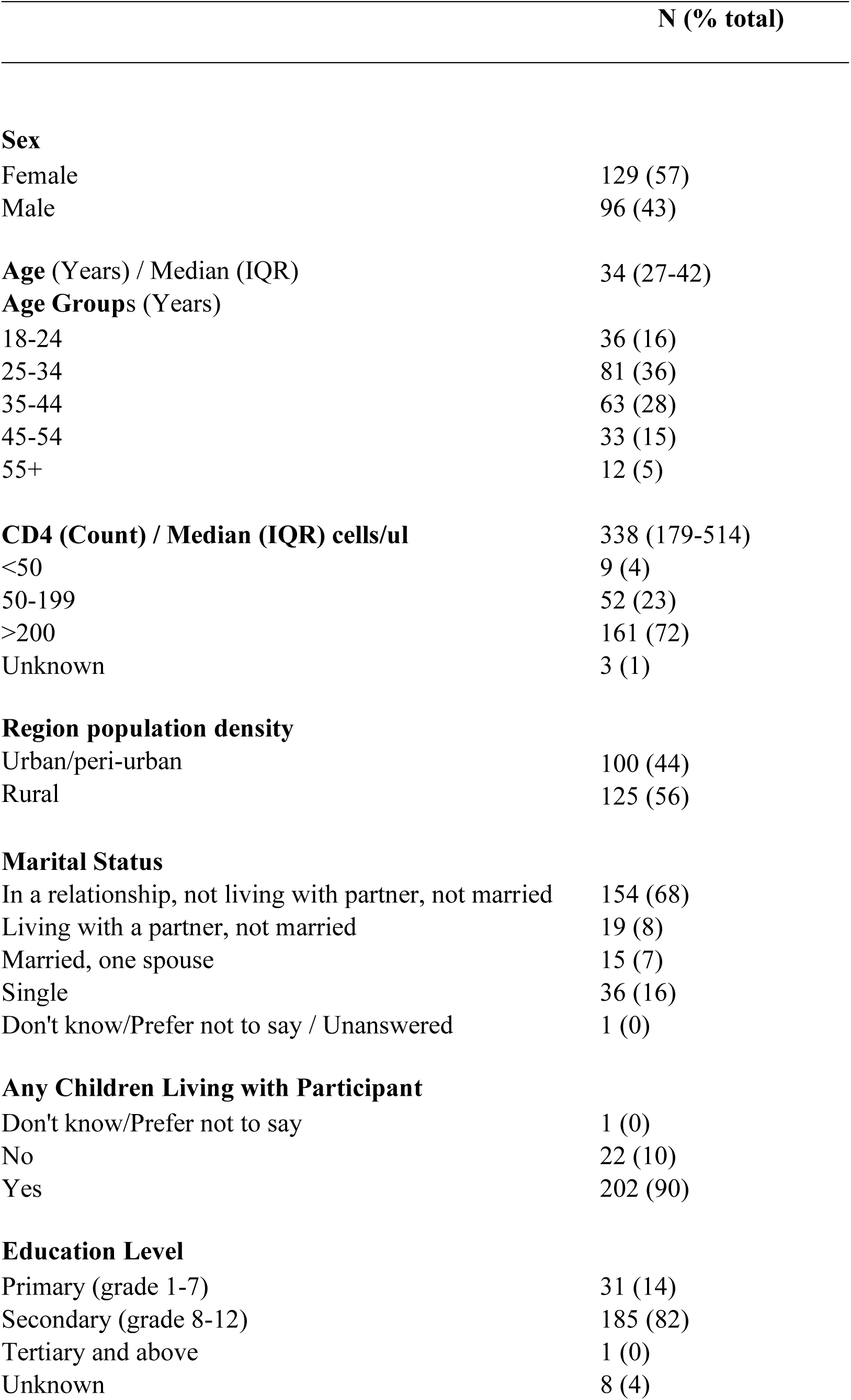
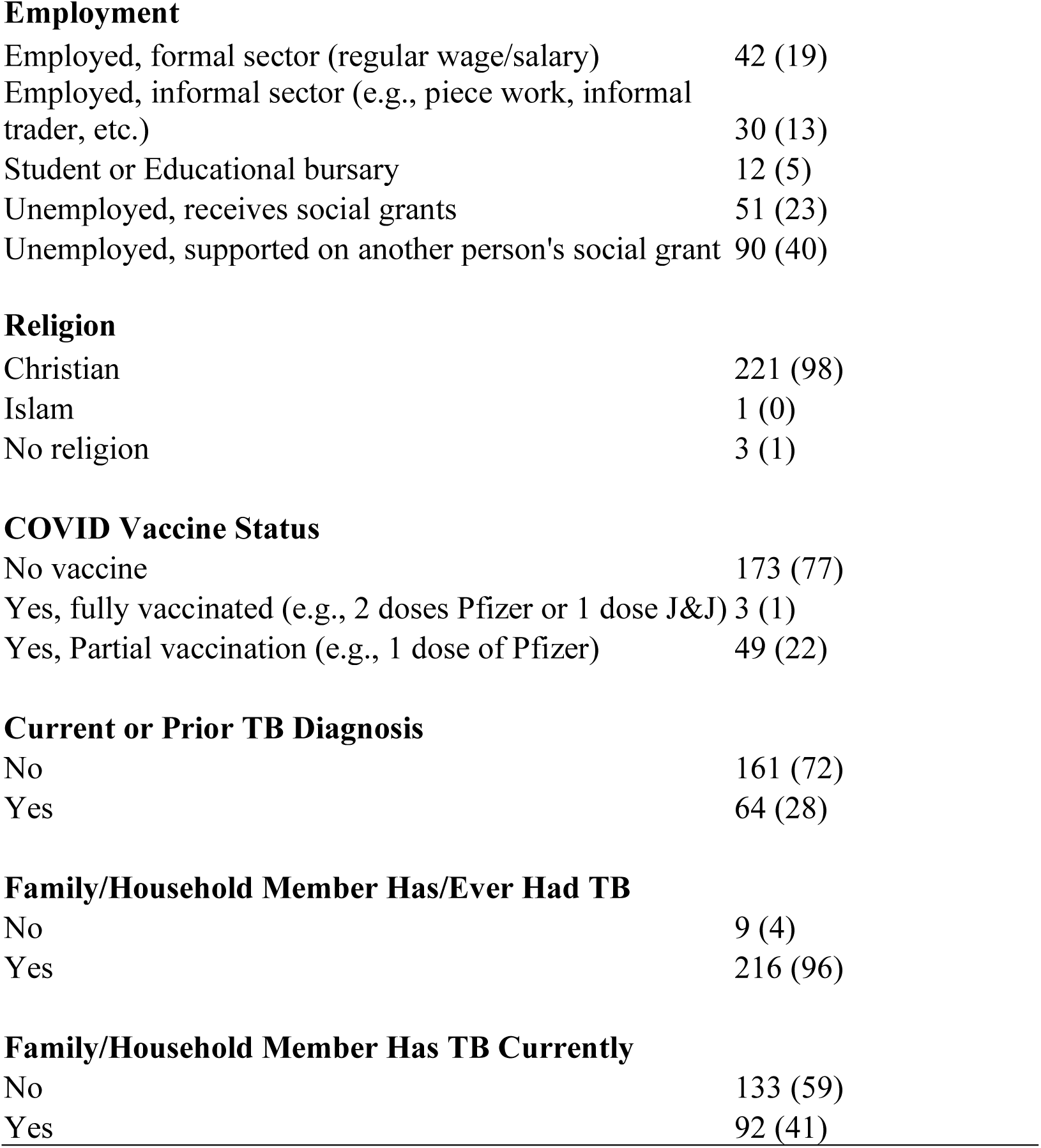
Clinical and demographic characteristics of respondents to the VAS.

All 225 (100%) participants felt TB is a somewhat, very, or extremely important problem in the community and it is important for scientists to find a new vaccine to prevent TB disease. All 225 (100%) reported they were somewhat, very, or extremely concerned about contracting TB; in contrast, any concern was lower for influenza (107, 48%), COVID-19 (61, 28%), malaria (6, 3%), or measles (50, 23%). All 225 (100%) were somewhat, very, or extremely concerned about a family member contracting TB, with lower concern for influenza (103, 57%), measles (114, 51%), COVID-19 (69, 31%), or malaria (27, 12%).

All 225 (100%) participants “strongly agreed” or “tended to agree” it was important for people of all ages to have vaccines and for scientists to discover a NTV. Similarly, 163 (72%) tended to agree and 60 (27%) strongly agreed a NTV is important. Figure 1 shows 181 (80%) participants tended to and 29 (13%) strongly agreed a NTV would be safe while 165 (73%) tended to agree and 56 (25%) strongly agreed a NTV would be effective. In response to questions about vaccines in general, 185 (82%) participants tended to agree and 24 (11%) strongly agreed that vaccines overall are safe, while 164 (73%) tended to agree and 54 (24%) strongly agreed vaccines overall are effective. Few participants disagreed, were unsure, or did not answer.

**Figure 1:**
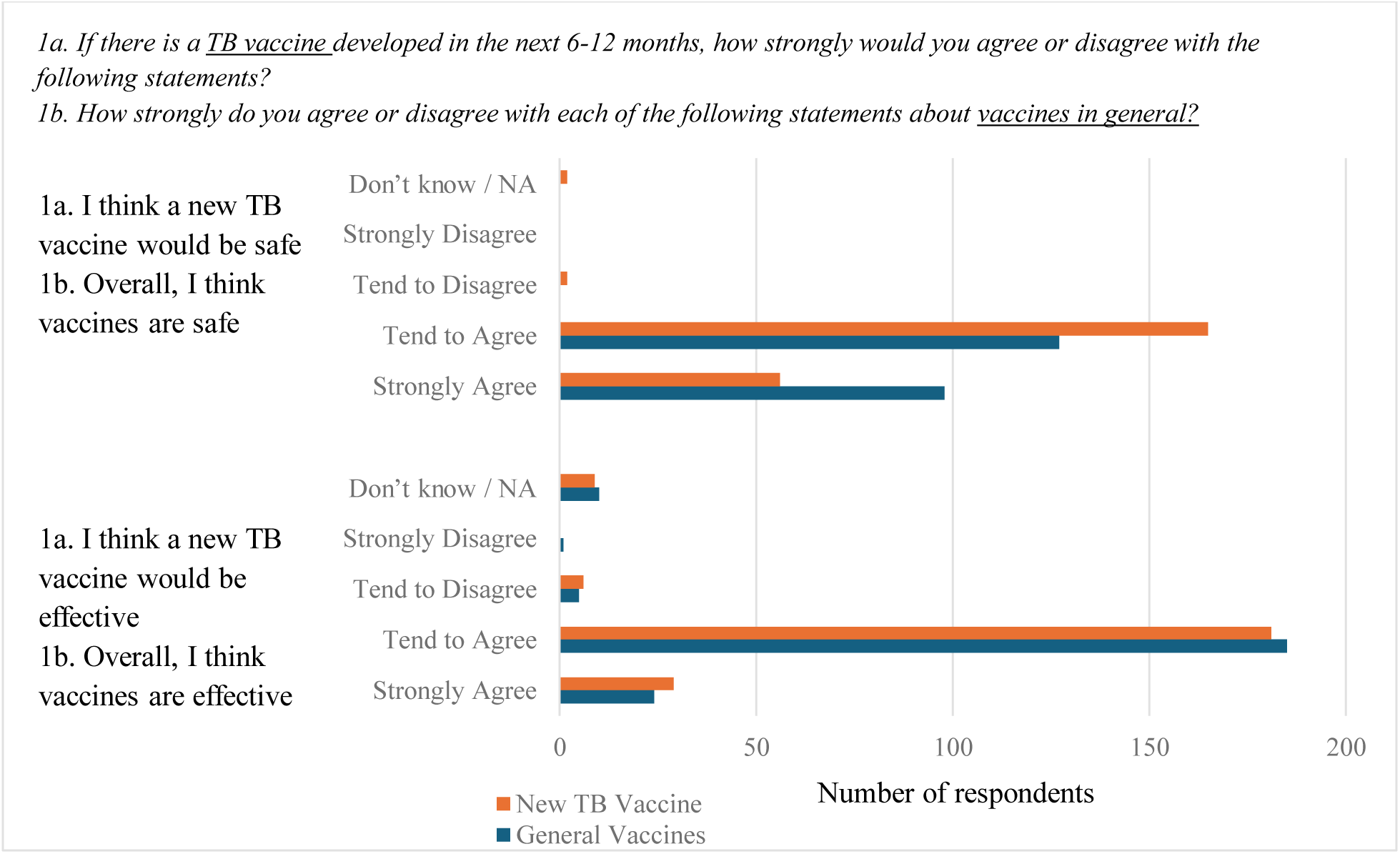
Confidence in the safety and effectiveness of a NTV and general vaccines. For willingness to receive a NTV, 198 (88%) responded “Definitely yes” and 27 (12%) responded that they were “Unsure, leaning yes,” with nobody responding, “leaning no” or “definitely no.” For timing of vaccination, 118 (52%) reported that they would take a NTV immediately, and 107 (48%) said that they would wait ≥6 months. Taken together, vaccine acceptability was reflected as 112 (50%) participants responding definitely yes, as soon as available to a NTV, 86 (38%) responding definitely yes, but wait at least six months, and 27 (12%) responding unsure, leaning towards yes, any timeline.

Participants were asked about willingness to receive a NTV of a specific platform type and were provided with examples of common vaccines for each platform. For a vaccine platform to prevent TB, responses of “definitely” or “unsure, leaning towards yes” were reported by 219 (98%) participants for a live attenuated vaccine like the BCG and by 200 (89%) participants for a protein subunit vaccine like hepatitis B. In contrast, responses of “definitely” or “leaning towards yes” were reported by only 90 (40%) participants for an mRNA vaccine like Pfizer or Moderna and 45 (20%) participants for a viral vector vaccine like J&J. All (225, 100%) were willing to talk to other people about getting a NTV. When asked if vaccines overall were compatible with their religion, 76 (34%) agreed, 87 (39%) tended to disagree, and 62 (28%) strongly disagreed.

Participants were asked about people and sources of information they would access for TB information and general health information (Figure 2). The most common sources of TB information were NGOs or community support organizations (202, 92%), a family doctor or personal health provider (201, 92%), government websites (190, 87%), or international health authorities (Africa CDC, WHO) (191, 87%). The most common sources of general health information were NGOs or community support organizations (215, 98%), family members (213, 97%), a family doctor or personal healthcare provider (213, 97%), government websites (208, 95%), traditional news like television and radio (207, 95%), international health authorities (203, 93%), other government communications online like social media (201, 92%), and online or hard copy newspapers (199, 91%). Of note, for both questions, few people (<5%) selected social media influencers or social media

**Figure 2:**
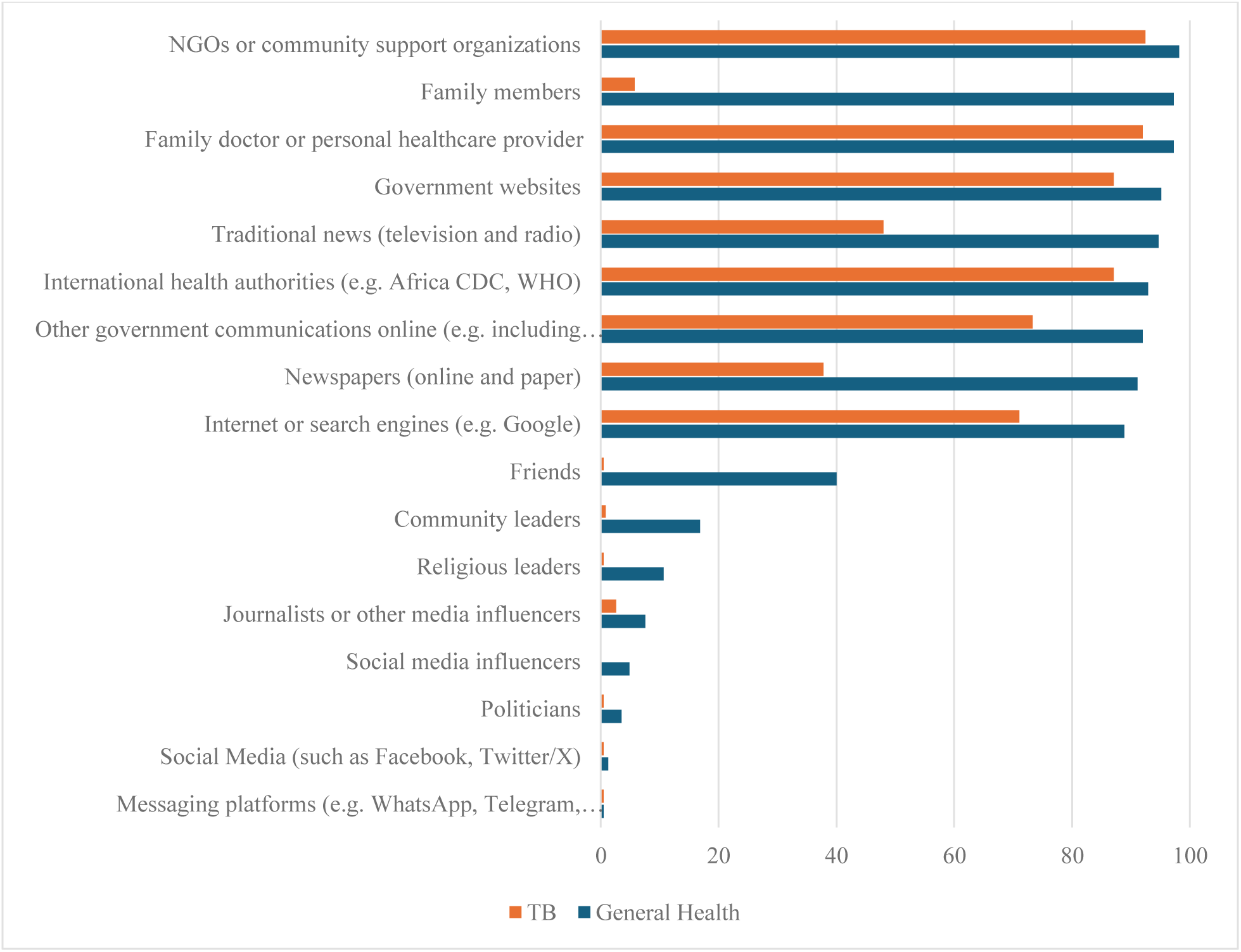
Information sources for general health and TB. Shown as % participants responding yes per category. Sources were asked independently, and multiple answers were permitted. Participants were asked, “Which of the following would you use to access news or information about: 1) Your general health and 2) Tuberculosis (TB)” and “Which of the following people / types of people would you turn to for information about: 1) Your general health and 2) Tuberculosis.”

Most participants reported a preference to receive a NTV in a community-based setting, such as a community center, meeting hall, local shop, or school (186, 85%) (Figure 3). Following this, 41 (18%) would prefer to receive the vaccine at a mobile vaccination center. Few preferred to obtain the vaccine at a health center / clinic (16, 7%), hospital (5, 2%), pharmacy (17, 8%), work (11, 5%), dedicated vaccination center (1, <1%), or place of worship (0, 0%).

**Figure 3:**
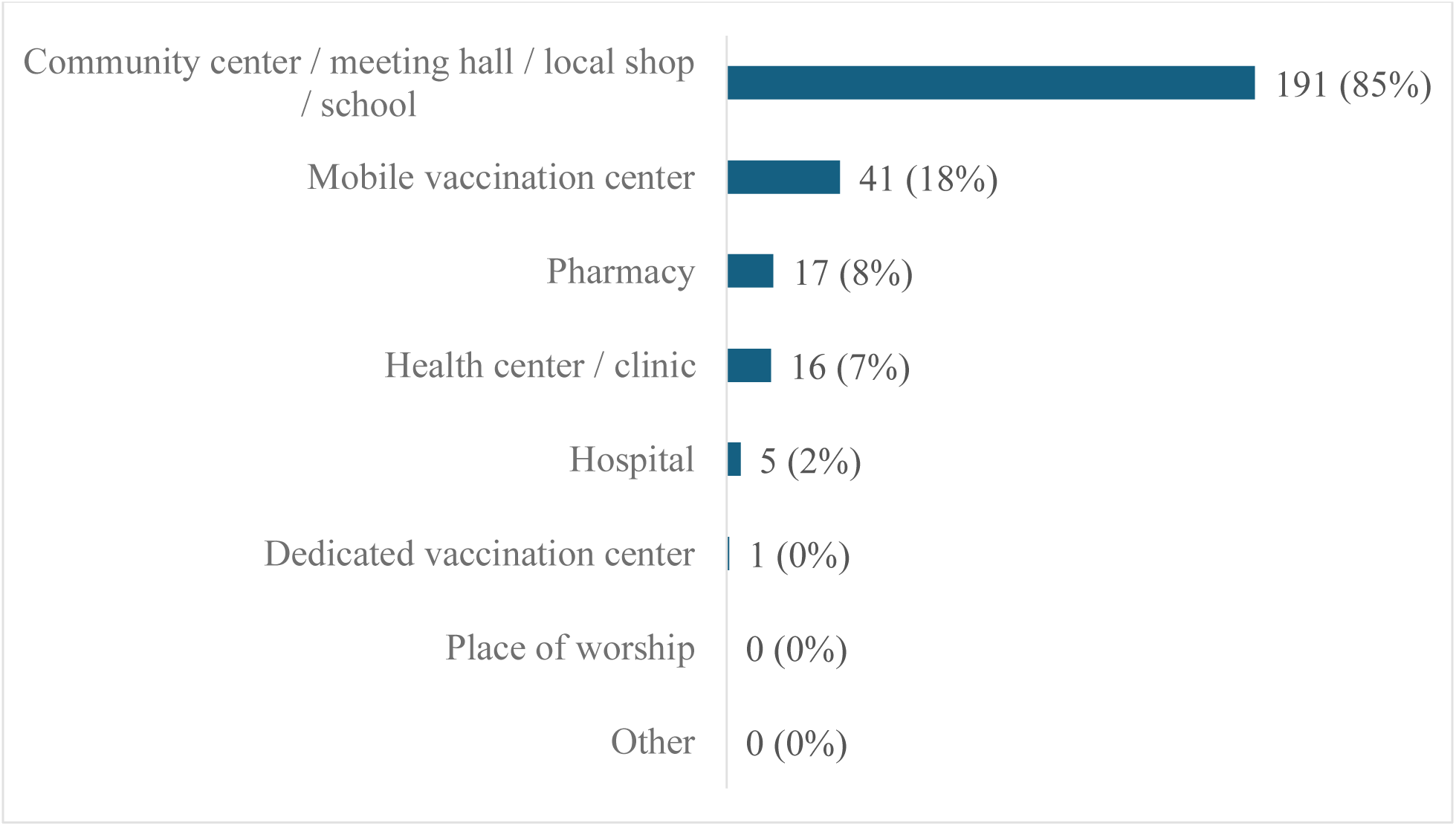
Preferred location to receive a NTV. Participants were asked, “If a new tuberculosis (TB) were available, WHERE would you prefer to get one? Please list as many locations as apply.”

Vaccine acceptability was observed to be significantly greater among participants who were extremely concerned about contracting TB (*p=*0.017), had a history of TB (*p=*0.044), perceived TB as an extremely important community health issue (p = 0.002), were unemployed (*p=*0.009), disagreed vaccines were compatible with their religion (*p<*0.001), or currently lived with someone with TB (*p=*0.044) (Figure 4). Age, sex, employment status, education level, geographic location, and COVID-19 vaccine status use were not significantly associated with NTV acceptability. Perception of vaccine importance and effectiveness were skewed towards participants tending to agree, and comparison of those strongly agreeing versus tending to agree within these categories was not significant. High skew in religion (98% Christian), social media use for TB information (1%), and perception of vaccine safety (>85% tending to agree) precluded meaningful analysis of these variables with vaccine acceptability.

**Figure 4:**
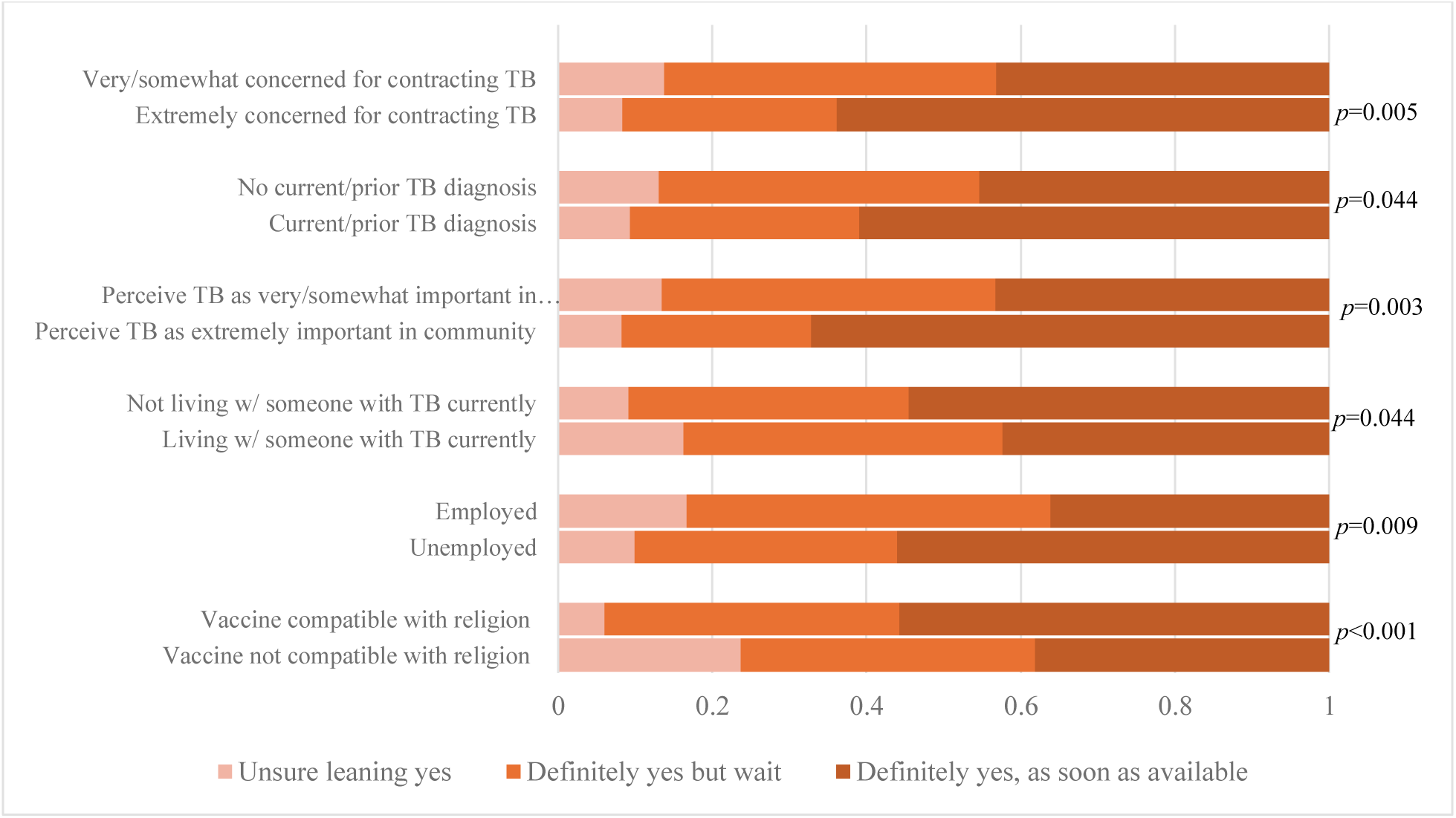
Sociodemographic factors and corresponding response levels that were significantly associated with acceptability of a NTV. The x-axis represents proportional distribution of vaccine acceptability within each sociodemographic group. Unless otherwise noted, all other variables and levels within each variable tested were not significantly associated with vaccine acceptability in this sample.

## Discussion

Acceptability and confidence in a NTV were high among this sample of South African PWH. Acceptability was observed to be greater among those who perceived TB to be an important community issue, had higher concern about contracting TB, had a history of TB, or were unemployed. There was an unexpected association between vaccine acceptability and reporting a belief that vaccines were incompatible with one’s religion, and a similarly surprising relationship of lower acceptability among people living with someone with TB. Respondents widely preferred to receive a NTV in a community space and preferred to receive general health or TB-specific information from health, government, and community authorities or the internet.

Comparison data for NTV acceptability is extremely limited. In a small sample of Mozambican adults and adolescents, 77% reported willingness to receive a NTV [22]. In a survey of 350 high-school students in Khayelitsha, South Africa, 84.1% would say yes to receiving a NTV [23] and in a sample of 499 adults in Zambia, the adjusted probability that community members would definitely get a NTV was 76.9% among the general community and 82.6% for healthcare workers [24]. Acceptability of COVID-19, HPV, and childhood immunizations in South Africa is lower [12,13,25]. When the COVID-19 vaccine was first introduced in late 2020, only 76% of South Africans reported willingness to accept it [11].

Confidence in overall and TB-specific vaccine safety and effectiveness was notably high in the study, precluding sufficient power to assess the relationship with vaccine acceptability. Previous studies have observed lower vaccine acceptability in individuals with concerns about vaccine safety, efficacy, or side effects in the contexts of COVID-19, HPV, and childhood immunizations [11,13–16].

Social desirability bias could have contributed to the high vaccine acceptability, confidence, and perception of importance seen in our study. However, concern about social desirability bias is mitigated by the willingness of participants to report high levels (77%) of non-vaccination against COVID-19. Additionally, while 28% of participants reported concern for contracting COVID-19, 100% reported concern for contracting TB. Taken together, vaccine acceptability seems driven by disease risk perception, which has also been demonstrated in Sub-Saharan African studies on HPV [15], COVID-19 [11,26], and TB [23,24]. Indeed, South Africa has one of the highest burdens of TB globally [1], and 96% of our participants reported ever living with someone with TB. Additionally, in 2023, 54% of TB cases in South Africa occurred in individuals living with HIV [17], so in our study population of exclusively PWH, the high perceived risk may have contributed to vaccine acceptability.

Our study did not find a consistent association between vaccine acceptability and gender [11,12,14,16], rurality [14,27], or education level [6,11,12,14], which is similar to others studies on COVID, HPV, and childhood immunizations. All study participants were Black, precluding analysis of an association between acceptability and race/ethnicity. In contrast to other studies, an association between NTV acceptability and older age was not observed [11–14,27,28]. This may be attributed to the more limited age range of our study participants, with only 12 participants being over 55 years old and no participants over 67 years old. A significant association was also found between NTV acceptability and unemployment, in contrast to acceptability studies of COVID-19 vaccines, which showed inconsistent or opposite relationships with unemployment [11,27].

We observed that participants who currently lived with someone with TB expressed less acceptability of NTVs. By contrast, not knowing anyone who tested positive for COVID was previously associated with being more hesitant to accept the COVID vaccine [11]. Participants who felt vaccines were incompatible with their religion reported greater vaccine acceptability. However, this finding should be interpreted with caution given small counts of individuals in the ‘Unsure, leaning toward yes’ categories. It has been previously described that compatibility of vaccines with religious beliefs is associated with greater COVID vaccine acceptability [6,12,14]. Lastly, despite 35% of rural and 20% of urban South Africans reporting use of social media as a trusted source for COVID-19 vaccine information [11], only one participant in this survey reported receiving health information from social media, precluding meaningful analysis. In the literature, receiving vaccine information from social media was strongly associated with COVID vaccine hesitancy [11,12]. Our findings are limited by small sample size or lack of qualitative context which may be clarified with more acceptability studies.

We observed strong preferences for certain vaccine platforms among respondents.

Participants were more interested in a future NTV that is like the BCG or hepatitis B than mRNA or viral vector vaccines used during COVID. This may be explained by participants’ greater familiarity with the BCG and hepatitis B vaccines which have been established in the South African childhood vaccination schedule since 1974 and 1995, respectively [29]. Indeed, there was public concern during the COVID-19 pandemic that testing and development of the COVID vaccines was too rushed [12].

While this study did not directly assess participant’s trust in health and government authorities, most respondents reported a preference to receive information on general health and TB specifically from community, health, and governmental entities or internet. Notably, participants were far less likely to seek TB information from family, friends, newspapers, or traditional news than they did for general health information. Participants did not frequently seek general or TB-specific health information from social media, journalists, media influencers, politicians, or religious leaders. These findings inform preferred communication and education platforms for the NTV in advance of its rollout. Future acceptability studies could also directly inquire about the significant association between vaccine acceptability and trust in government [6,14,16] or health care providers [14,25] previously seen with COVID, HPV, and childhood immunizations.

Finally, this study identified participants’ strong preference to receive NTVs in a community space rather than a dedicated health center, which has implications for selecting settings for future NTV campaigns and delivery. Additional context from qualitative interviews and other methods would help identify reasons for this preference, including the contributions of concerns about health care accessibility, stigma, or other reasons.

## Strengths and Limitations

Strengths of the study include the highly relevant sample population for NTV. PWH in South Africa represent a priority recipient population for a NTV given the high prevalence and mortality rate of TB among PWH and high burden of TB in South Africa [2,17]. The sample populations captured two geographic settings, rural and urban, which improves the generalizability of findings in South Africa. The study fills a critical gap in NTV acceptability studies to date, which are time-sensitive given WHO’s goal of introducing a NTV by 2030.

This analysis had multiple comparisons, therefore, observed associations must be interpreted with caution as they are not designed for prediction. The use of univariable Kruskal-Wallis analyses did not account for potentially interacting variables. Equally, the small sample size may have limited statistical power, and therefore the precision of associations found. The quantitative nature of the study did not capture context behind unexpected findings, such as the association between vaccine acceptability and incompatibility of vaccines with religion or living with someone with TB. The study cohort only included clinic-attending adults living with HIV so findings may not be generalizable to the entire population of potentially vaccine-eligible persons including adolescents, people without HIV, or people not in clinical care. Data was collected in one province of South Africa which may limit generalizability to other vaccine-eligible settings like Asia and Latin America where the TB epidemic is less HIV-driven.

## Conclusions

Several NTVs are in clinical trials, but community perceptions are poorly characterized and urgently needed to inform demand creation among global public health stakeholders. This study found that acceptability of and confidence in NTVs were high among a sample of South African adults living with HIV. Acceptability was observed to be associated with perceived risk of TB disease, perceived importance of TB as a health problem in the community, and unemployment. Additionally, participants preferred to receive NTVs in a community space and receive TB information from government, health, and community entities and internet but not from traditional news (TV, radio, newspaper), friends, family, or social media. These findings may inform public communication strategies, information platforms, and settings for vaccine administration ahead of NTV campaigns.

## Data Availability

All data produced in the present study are available upon reasonable request to the authors

## Declaration of Interest Statement

□ The authors declare that they have no known competing financial interests or personal relationships that could have appeared to influence the work reported in this paper.
□ The author is an Editorial Board Member/Editor-in-Chief/Associate Editor/Guest Editor for this journal and was not involved in the editorial review or the decision to publish this article.
☒ The authors declare the following financial interests/personal relationships which may be considered as potential competing interests:

Adrienne E. Shapiro, MD, PhD reports receiving grants to her institution from Merck and Gilead as a clinical trial investigator and receiving consulting fees from Merck; Jennifer M. Ross, MD, MPH reports receiving research grants to her institution from Merck. All other authors report no known competing financial interests or personal relationships that could have appeared to influence the work reported in this paper.

## Declaration of generative AI in scientific writing

During the preparation of this work, Alexa Giovanatti used ChatGPT to help guide statistical analysis, data interpretation, and writing code. After using this tool/service, the author(s) reviewed and edited the content as needed and take(s) full responsibility for the content of the published article.

## Acknowledgements

The authors thank the DROP-TB study staff, Umkhanyakude District Clinic Staff, and the UW/Fred Hutch CFAR Biostatistics Consultation Service in their support of the study execution.

## Ethical Approval

The study was approved by the University of Washington Institutional Review Board (Study 00006518), the University of KwaZulu-Natal Biomedical Research Ethics Committee (BREC/00001174/2020), and the London School of Hygiene and Tropical Medicine Ethics Committee (Ref. 21624). The study was also approved by the Health Research Committee of the KwaZulu-Natal Department of Health (NHRD KZ_202010_029).

## Supplement

### Survey

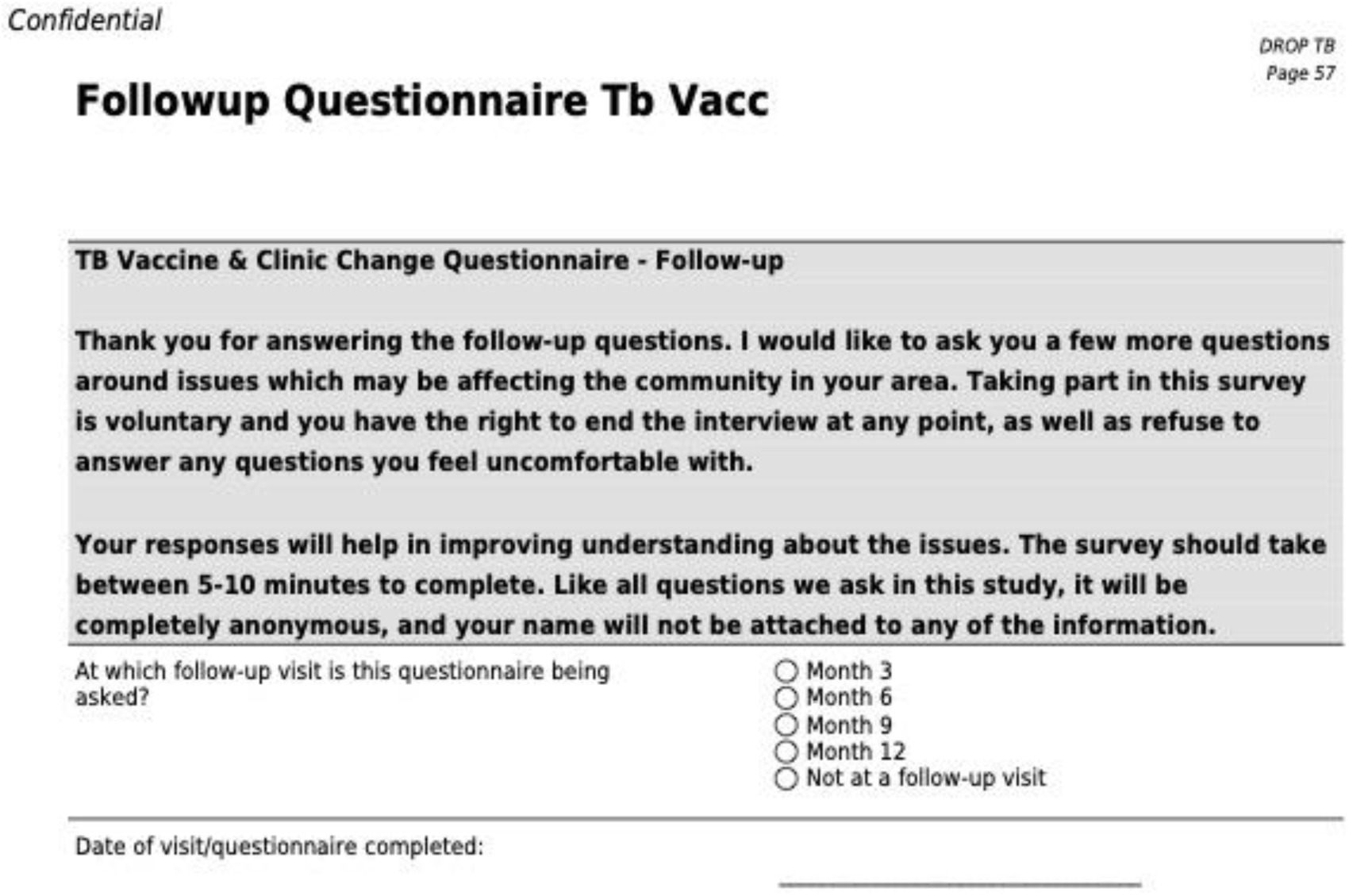
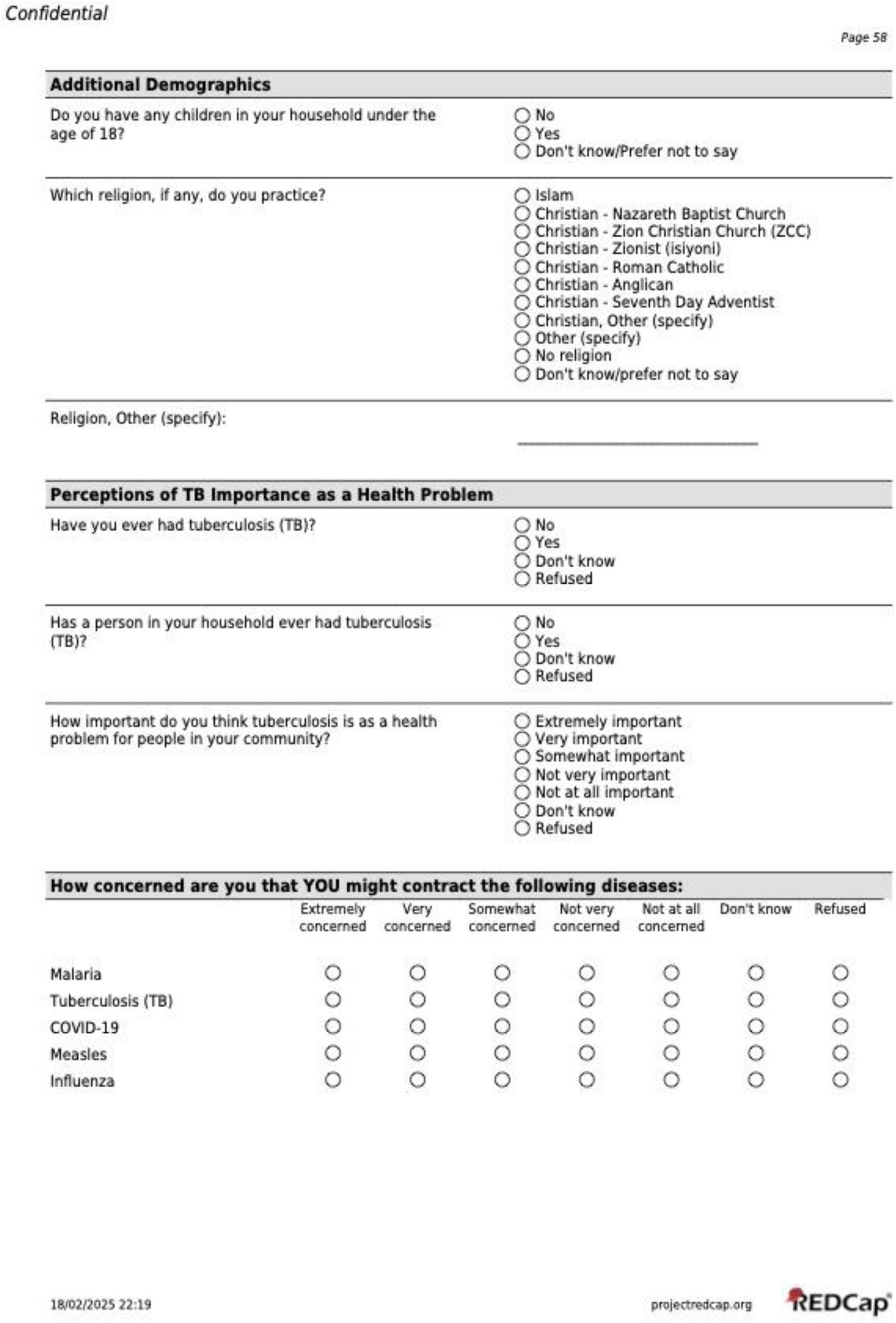
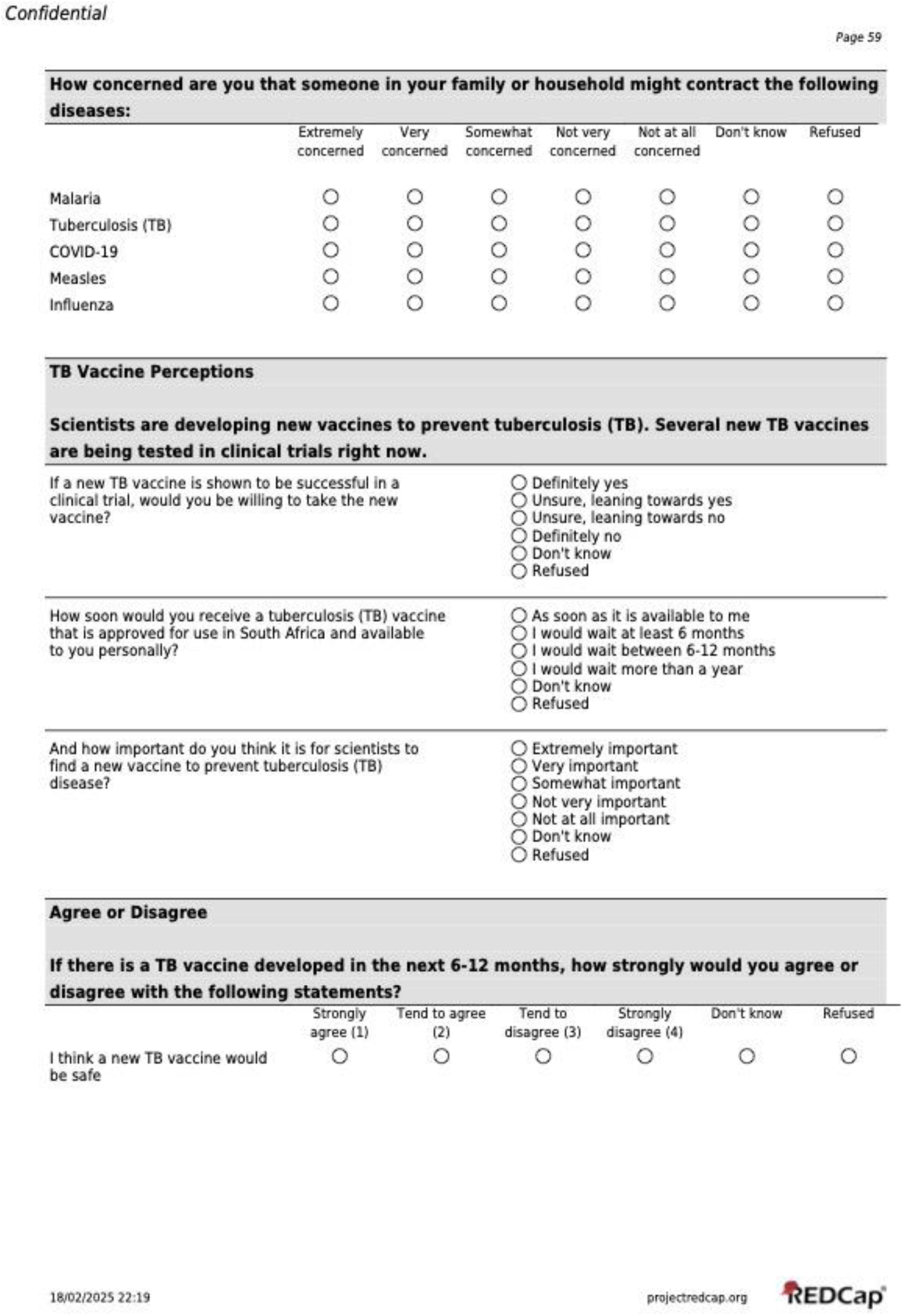
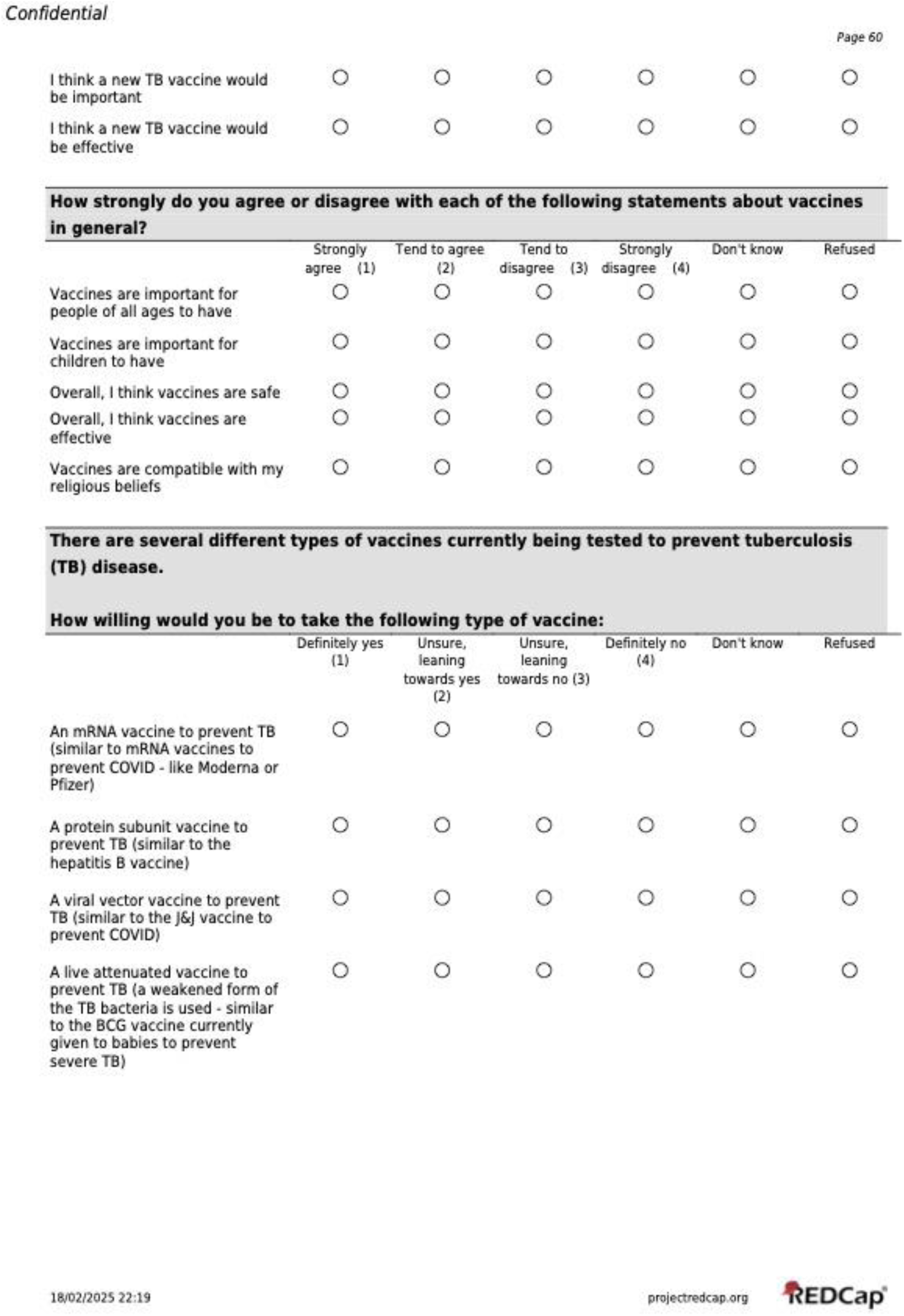
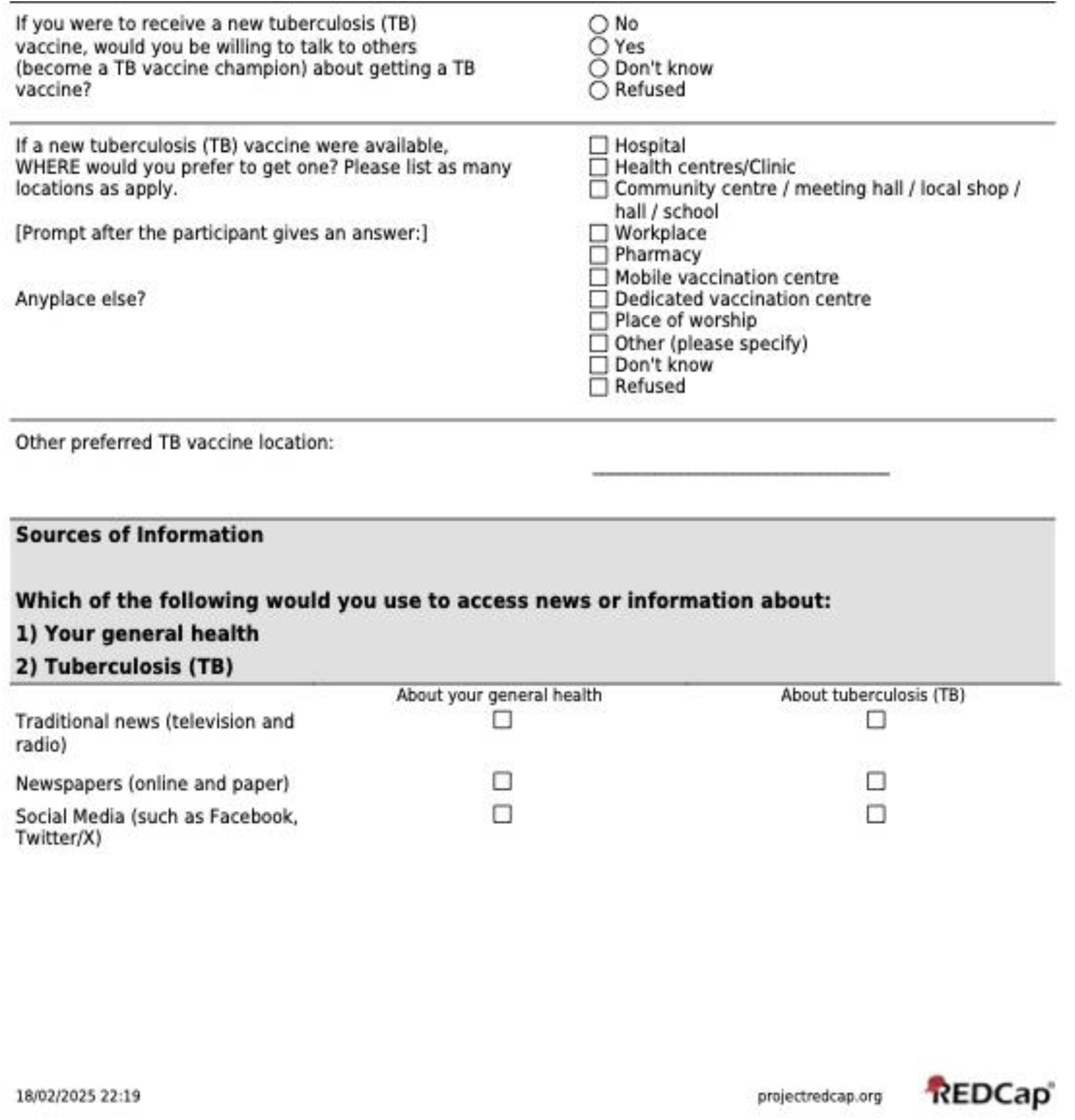
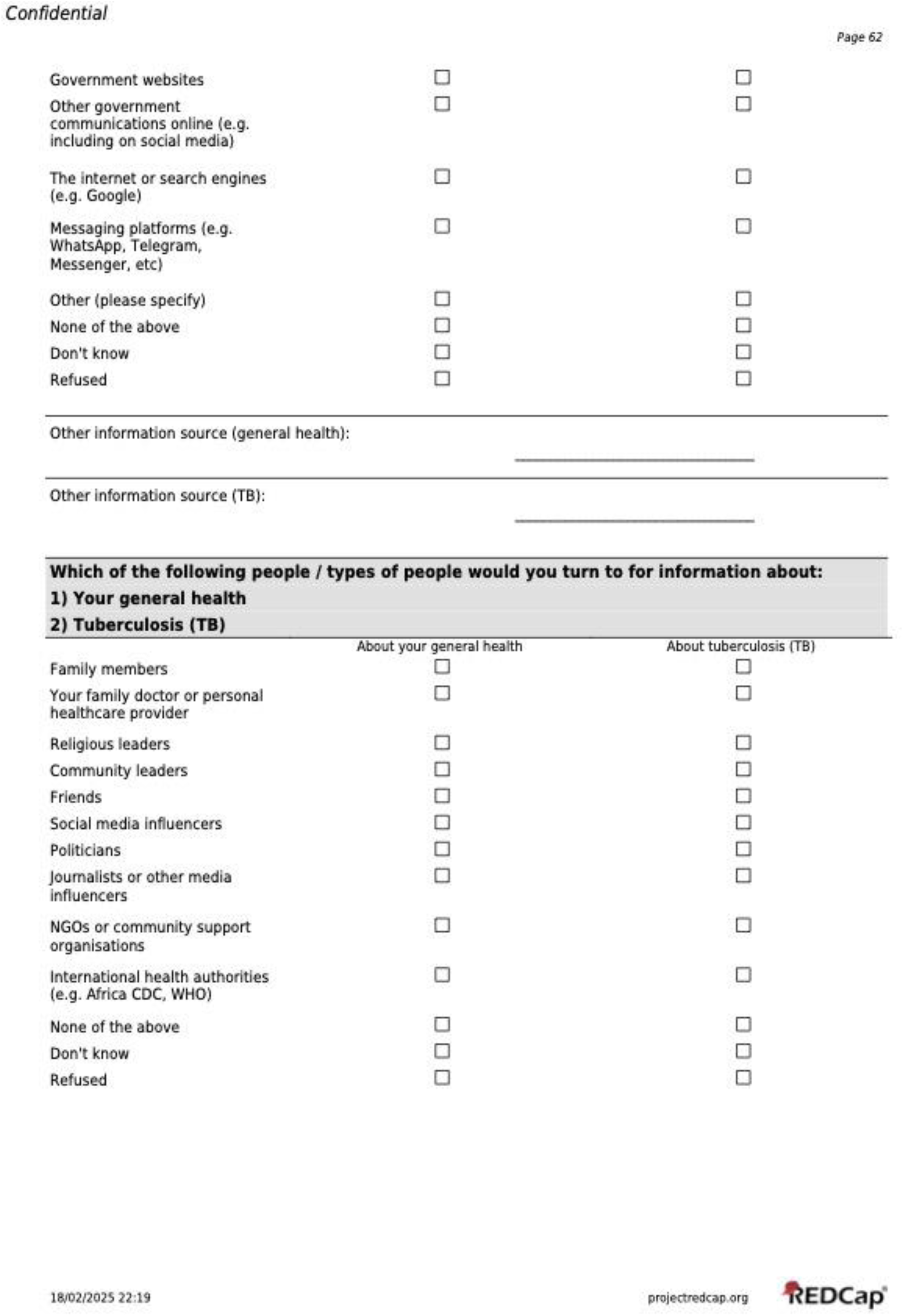
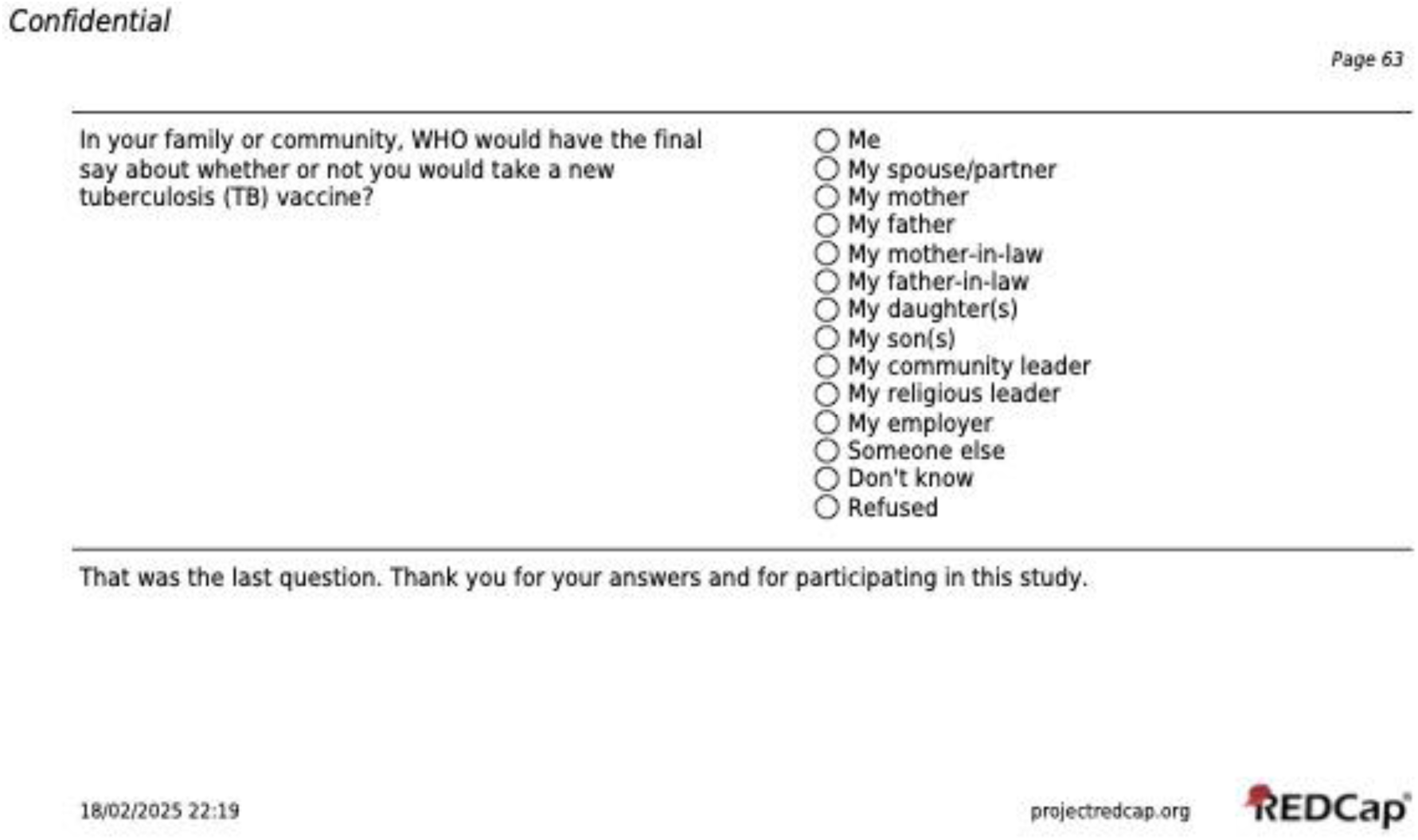

### Statistical analyses

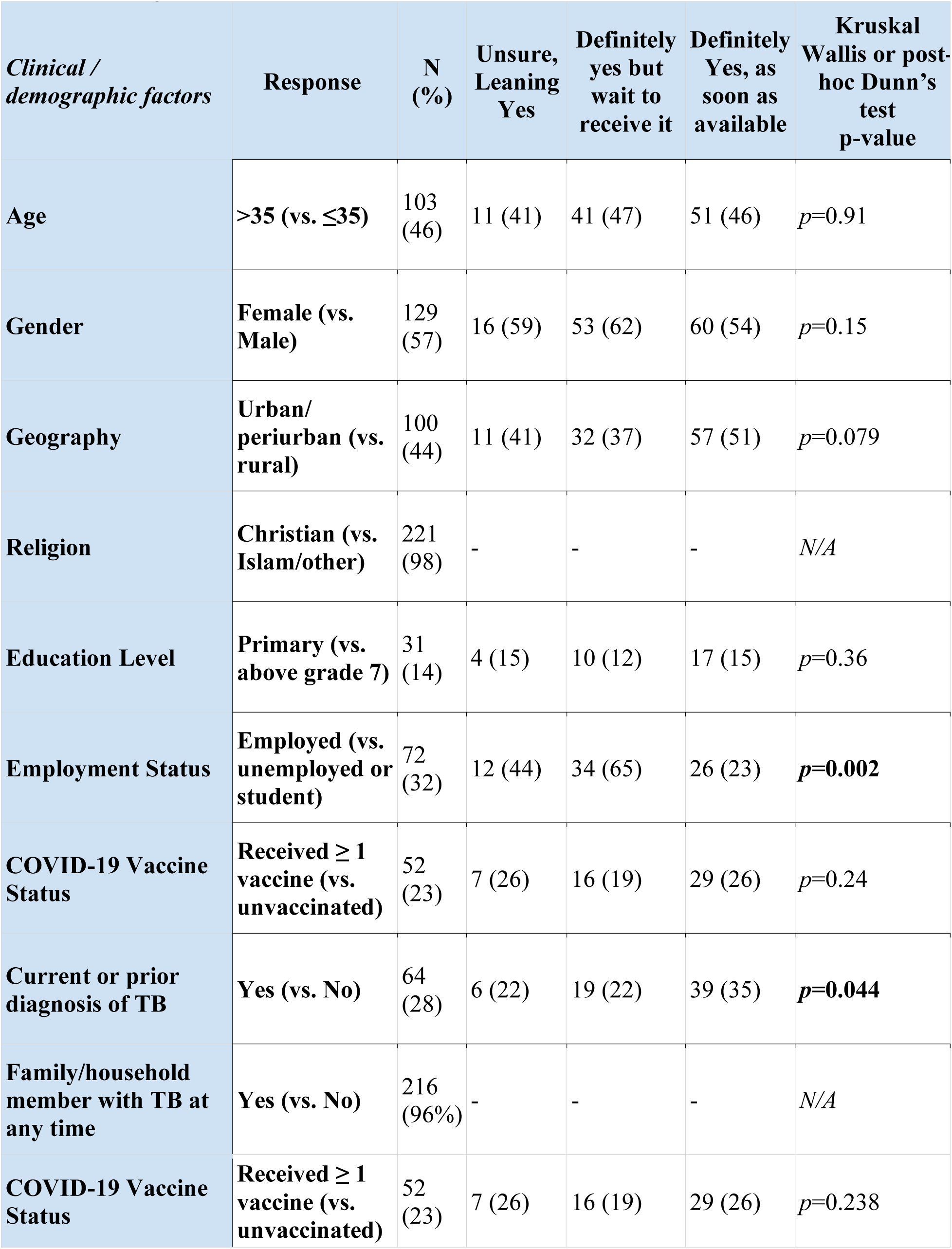
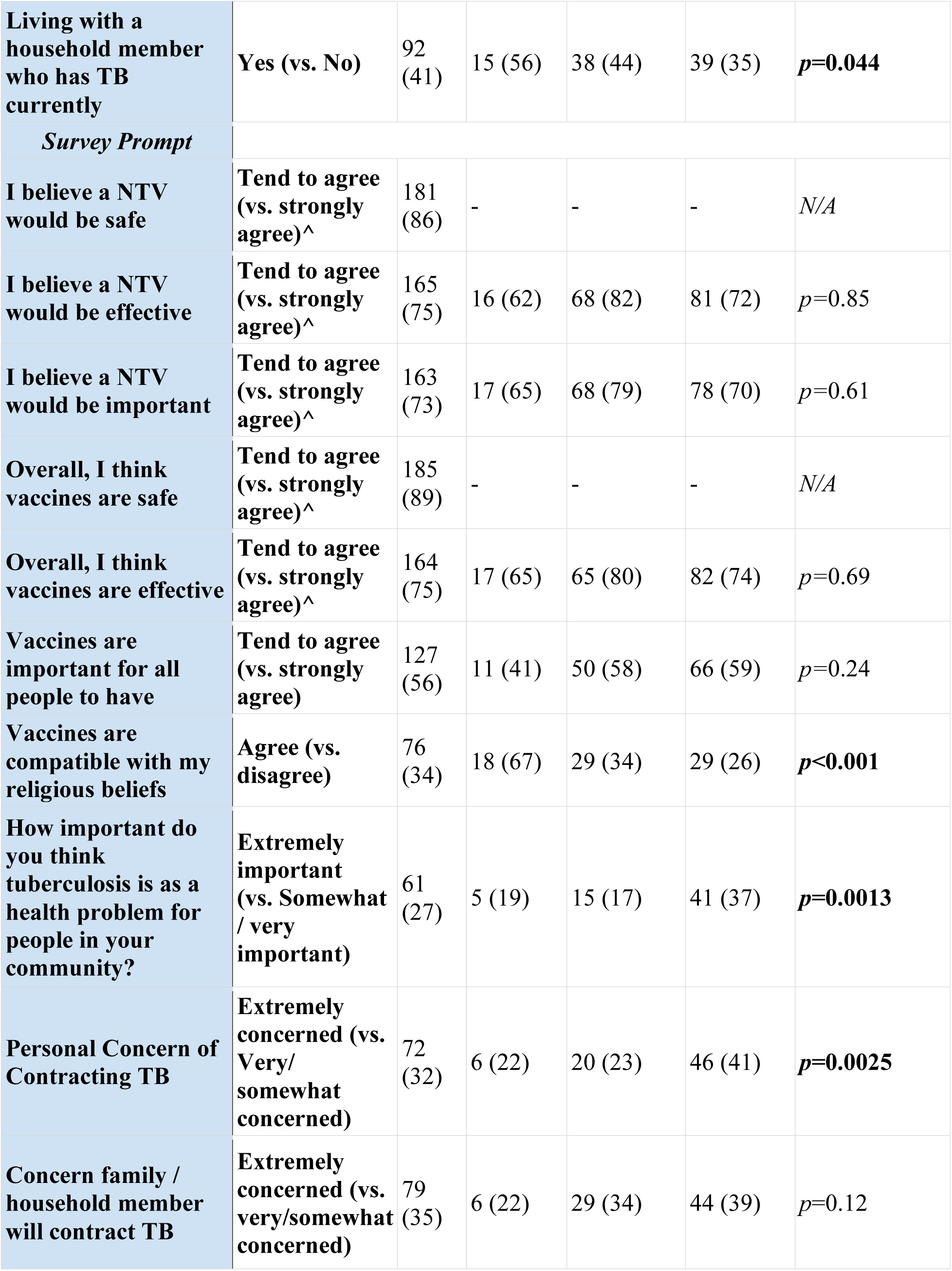
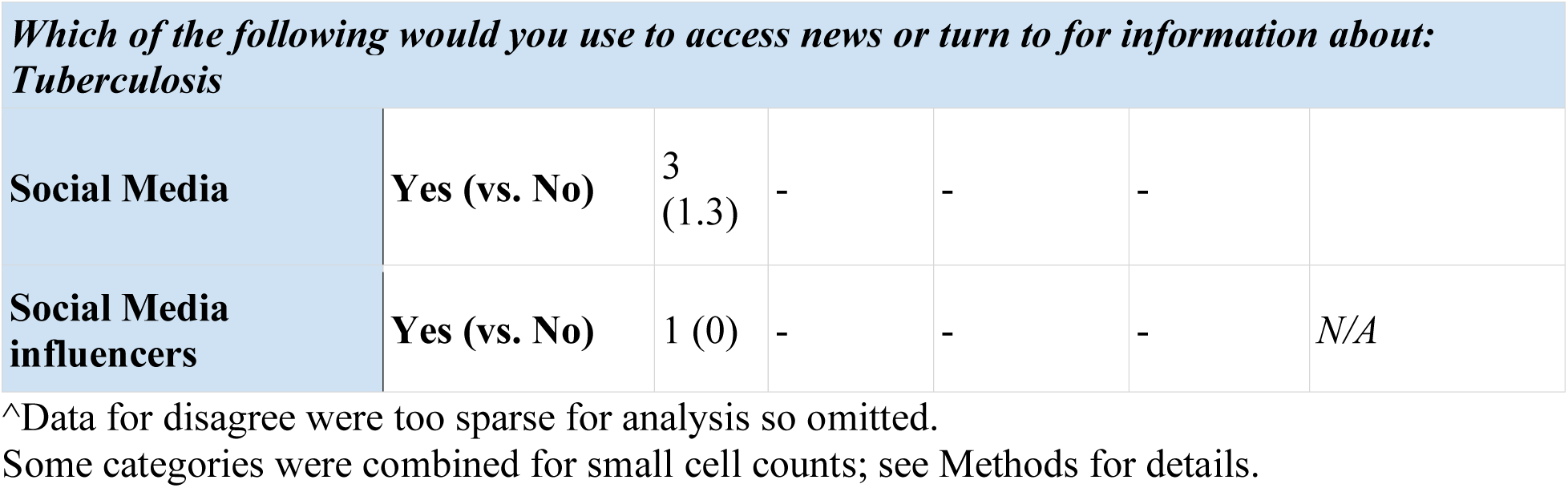

| *Clinical / demographic factors* | Response | N (%) | Unsure, Leaning Yes | Definitely yes but wait to receive it | Definitely Yes, as soon as available | Kruskal Wallis or post-hoc Dunn's test p-value |
| --- | --- | --- | --- | --- | --- | --- |
| Age | >35 (vs. ≤35) | 103 (46) | 11 (41) | 41 (47) | 51 (46) | $p=0.91$ |
| Gender | Female (vs. Male) | 129 (57) | 16 (59) | 53 (62) | 60 (54) | $p=0.15$ |
| Geography | Urban/periurban (vs. rural) | 100 (44) | 11 (41) | 32 (37) | 57 (51) | $p=0.079$ |
| Religion | Christian (vs. Islam/other) | 221 (98) | - | - | - | $N/A$ |
| Education Level | Primary (vs. above grade 7) | 31 (14) | 4 (15) | 10 (12) | 17 (15) | $p=0.36$ |
| Employment Status | Employed (vs. unemployed or student) | 72 (32) | 12 (44) | 34 (65) | 26 (23) | $p=0.002$ |
| COVID-19 Vaccine Status | Received ≥ 1 vaccine (vs. unvaccinated) | 52 (23) | 7 (26) | 16 (19) | 29 (26) | $p=0.24$ |
| Current or prior diagnosis of TB | Yes (vs. No) | 64 (28) | 6 (22) | 19 (22) | 39 (35) | $p=0.044$ |
| Family/household member with TB at any time | Yes (vs. No) | 216 (96%) | - | - | - | $N/A$ |
| COVID-19 Vaccine Status | Received ≥ 1 vaccine (vs. unvaccinated) | 52 (23) | 7 (26) | 16 (19) | 29 (26) | $p=0.238$ |
| **Living with a household member who has TB currently** | **Yes (vs. No)** | 92 (41) | 15 (56) | 38 (44) | 39 (35) | ***p*=0.044** |
| *Survey Prompt* |  |  |  |  |  |  |
| **I believe a NTV would be safe** | **Tend to agree (vs. strongly agree)^** | 181 (86) | - | - | - | *N/A* |
| **I believe a NTV would be effective** | **Tend to agree (vs. strongly agree)^** | 165 (75) | 16 (62) | 68 (82) | 81 (72) | *p*=0.85 |
| **I believe a NTV would be important** | **Tend to agree (vs. strongly agree)^** | 163 (73) | 17 (65) | 68 (79) | 78 (70) | *p*=0.61 |
| **Overall, I think vaccines are safe** | **Tend to agree (vs. strongly agree)^** | 185 (89) | - | - | - | *N/A* |
| **Overall, I think vaccines are effective** | **Tend to agree (vs. strongly agree)^** | 164 (75) | 17 (65) | 65 (80) | 82 (74) | *p*=0.69 |
| **Vaccines are important for all people to have** | **Tend to agree (vs. strongly agree)** | 127 (56) | 11 (41) | 50 (58) | 66 (59) | *p*=0.24 |
| **Vaccines are compatible with my religious beliefs** | **Agree (vs. disagree)** | 76 (34) | 18 (67) | 29 (34) | 29 (26) | ***p*<0.001** |
| **How important do you think tuberculosis is as a health problem for people in your community?** | **Extremely important (vs. Somewhat / very important)** | 61 (27) | 5 (19) | 15 (17) | 41 (37) | ***p*=0.0013** |
| **Personal Concern of Contracting TB** | **Extremely concerned (vs. Very/ somewhat concerned)** | 72 (32) | 6 (22) | 20 (23) | 46 (41) | ***p*=0.0025** |
| **Concern family / household member will contract TB** | **Extremely concerned (vs. very/somewhat concerned)** | 79 (35) | 6 (22) | 29 (34) | 44 (39) | *p*=0.12 |
| ***Which of the following would you use to access news or turn to for information about: Tuberculosis*** |  |  |  |  |  |  |
| **Social Media** | **Yes (vs. No)** | 3<br>(1.3) | - | - | - |  |
| **Social Media influencers** | **Yes (vs. No)** | 1 (0) | - | - | - | *N/A* |

^Data for disagree were too sparse for analysis so omitted.
Some categories were combined for small cell counts; see Methods for details.

